# Associations between Five-Year Blood Pressure Variability and Risk of Cardiovascular Events and Mortality

**DOI:** 10.1101/2020.12.21.20248682

**Authors:** Jiandong Zhou, Sharen Lee, Wing Tak Wong, William KK Wu, Wai Kit Ming, Tong Liu, Kamalan Jeevaratnam, Bernard Man Yung Cheung, Gary Tse, Qingpeng Zhang

**Affiliations:** School of Data Science, City University of Hong Kong, Hong Kong, Hong Kong SAR, China; Laboratory of Cardiovascular Physiology, Li Ka Shing Institute of Health Sciences, China; School of Life Sciences, The Chinese University of Hong Kong, Hong Kong SAR, China; Department of Anaesthesia and Intensive Care, Li Ka Shing Institute of Health Sciences, Hong Kong SAR, China; Department of Public Health and Preventive Medicine, School of Medicine, Jinan University, Guangzhou, China; Tianjin Key Laboratory of Ionic-Molecular Function of Cardiovascular disease, Department of Cardiology, Tianjin Institute of Cardiology, Second Hospital of Tianjin Medical University, Tianjin 300211, China; Faculty of Health and Medical Sciences, University of Surrey, GU2 7AL, Guildford, United Kingdom; Division of Clinical Pharmacology and Therapeutics, Department of Medicine, The University of Hong Kong, Pokfulam, Hong Kong SAR, China

## Abstract

**Introduction:** Blood pressure variability, in addition to blood pressure itself, has been used as a predictor for mortality. This study examined the predictive power of baseline/latest/mean/median blood pressure and blood pressure variability measures for all-cause mortality and adverse cardiovascular outcomes.

**Methods:** The retrospective observational study analyzed patients who presented to family medicine clinics between 1^st^ January, 2000 and 31^st^ December, 2001. Blood pressure measurements were obtained over a five-year period. Standard deviation (SD), root mean square (RMS), coefficient of variation (CV) and a variability score (number of >=5 mmHg blood pressure change) were used as measures of blood pressure variability. The primary outcome was all-cause mortality and the secondary outcomes were heart failure, acute myocardial infarction, and transient ischemic attack (TIA)/stroke, with follow-up until 31 December 2019.

**Results:** This study included 37540 patients (n=29597 patients with >=3 blood pressure measurements). A nonlinear inverse U-shaped relationship was observed between baseline/latest/maximum/minimum/mean/median/RMS measures of diastolic blood pressure and time-to-death for all-cause mortality (P<0.001). Higher variance/SD/CV/variability score of both systolic and diastolic blood pressure was significantly associated with increased risks of all-cause mortality and heart failure, acute myocardial infarction and TIA/stroke (P<0.001). Low baseline/latest/maximum/minimum/mean/median/RMS systolic blood pressure was significantly associated with shorter time-to-death for all-cause mortality (P<0.001).

**Conclusion:** Nonlinear inverse U-shaped relationships were observed between blood pressure and its variability measures and all-cause mortality. Higher blood pressure variability was associated with increased risk of all-cause mortality, heart failure, acute myocardial infarction and TIA/stroke.

## Introduction

Blood pressure variability, in addition to blood pressure itself, can be used as a predictor for adverse cardiovascular events and mortality in various disease cohorts including diabetes and hypertension ^1-3^. This can be assessed continuously over a period of time or can be determined as between visits to the clinic or hospital ^4^. There are several methods that can be used to calculate variability, such as standard deviation (SD) or coefficient of variation (CV). However, a score based on the frequency exceeding a fixed change in the absolute values has not been examined for blood pressure, in contrast to Hba1c variability. In this study, we systematically compared different blood pressure variability measures for their ability to predict all-cause mortality and adverse cardiovascular outcomes of heart failure, acute myocardial infarction and transient ischaemic attack (TIA)/stroke in patients attending family medicine clinics in Hong Kong.

## Methods

### Study Population

The study was approved by The Joint Chinese University of Hong Kong – New Territories East Cluster Clinical Research Ethics Committee and Institutional Review Board of the University of Hong Kong/Hospital Authority Hong Kong West Cluster. The inclusion criteria were patients who attended family medicine clinics in the Hong Kong Hospital Authority between 1^st^ January 2000 to 31^st^ December 2001. This made use of the Clinical Data Analysis and Reporting System (CDARS), a healthcare database that integrates patient information to establish comprehensive medical records with accurately linked mortality data. The system has been used for epidemiological research by multiple research teams, including our team, in the past ^5, 6^.

### Data extraction

Clinical and biochemical data were extracted for the present study. Patients demographics include sex and age of first blood pressure measurement. Prior comorbidities before first blood pressure tests are extracted, including cardiovascular, respiratory, kidney, endocrine, diabetes mellitus, hypertension, gastrointestinal diseases, based on the comorbidities data obtained before or on the first clinic attendance. Mortality data were obtained to the local death registry. The secondary outcomes and comorbidities were documented in CDARS under International Classification of Diseases (ICD)-9 codes. **Supplementary Appendix** displays the ICD codes used to search for patient outcomes and comorbidities. Cardiovascular and diabetes medications prescribed for the cohort patients were extracted.

Baseline biochemical data, defined as complete blood count, liver and renal function tests, diabetes mellitus and lipid tests measured from January 1^st^, 2000 to December 31^st^, 2001, were extracted. Complete blood count includes: mean corpuscular volume (MCV), fL; basophil, ×10^9/L; eosinophil, ×10^9/L; lymphocyte, ×10^9/L; blast, ×10^9/L; metamyelocyte, ×10^9/L; monocyte, ×10^9/L; neutrophil, ×10^9/L; white blood count, ×10^9/L; mean corpuscular hemoglobin (MCH), pg; myelocyte, ×10^9/L; platelet, ×10^9/L; reticulocyte, ×10^9/L; red blood count, ×10^12/L; and hematocrit (HCT), L/L. Liver and renal function tests include K/potassium, mmol/L; urate, mmol/L; albumin, g/L; Na/sodium, mmol/L; urea, mmol/L; protein, g/L; creatinine, μmol/L; alkaline phosphatase (ALP), U/L; aspartate transaminase, U/L; serum alanine aminotransferase (ALT), U/L; and bilirubin, μmol/L. Diabetes mellitus and lipid tests include HbA1c, %; cholesterol, mmol/L; glucose, mmol/L; and triglycerides, mmol/L.

### Variability calculations

Patients with three or more blood pressure measurements were included in the analysis for blood pressure variability. To calculate blood variability, data points were obtained for the five-year period between 1^st^ January 2000 and 31^st^ December 2004 using 1) standard deviation (SD), 2) coefficient of variation given by SD / mean, 3) variability score defined as 100 × no. of measurements > 5 mmHg / no. of measurements

### Outcomes and statistical analyses

The primary outcome was all-cause mortality. The secondary outcomes were heart failure, acute myocardial infarction and transient ischemic attack (TIA)/stroke. Follow-up was until 31^st^ December 2019. Descriptive statistics were used to summarize patients’ characteristics of primary outcome and secondary outcomes of incident heart failure, acute myocardial infarction and TIA/stroke. Continuous variables were presented as median (95% confidence interval [CI] or interquartile range [IQR]) and categorical variables were presented as frequency (%). The Mann-Whitney U test was used to compare continuous variables. The χ^2^ test with Yates’ correction was used for 2×2 contingency data, and Pearson’s χ^2^ test was used for contingency data for variables with more than two categories. Baseline/maximum/minimum values of diastolic and systolic blood pressure are extracted, and the temporal variability of diastolic and systolic blood pressure was examined through the following approaches: 1) mean, 2) median, 3) standard deviation (SD), 4) root mean square (RMS) by first squaring all blood pressure values then performing square root of the mean of the squares, 5) coefficient of variation (CV) by dividing the blood pressure standard deviation by the mean blood pressure then multiplying by 100, and 6) a variability score (from 0 [low] to 100 [high]) defined as the number of changes in blood pressure of 5 mmHg or more. To evaluate the significant prognostic risk factors and the effects of blood pressure and its variability measures associated with disease group status and primary outcomes, univariate Cox regression models was used with adjustments based on baseline characteristics. Time-to-death was defined by the number of months from first blood pressure examination to the date of mortality of the patient, or until December 31, 2019. Hazard ratios (HRs) with corresponding 95% confidence intervals (Cis) and P-values were reported accordingly. In addition, the associations between baseline/latest/mean/median of blood pressure and time-to-death for all-cause mortality was modeled using the generalized additive model, with 95% CI displayed. Kaplan-Meier curves were plotted against the time-to-death for mortality, incident heart failure, acute myocardial infarction and TIA/stroke, and stratified by patient sex. All significance tests were two-tailed and considered statistically significant when P-values <0.05. Data analyses were performed using RStudio software (Version: 1.1.456) and Python (Version: 3.6). Experiments were simulated on a 15-inch MacBook Pro with 2.2 GHz Intel Core i7 Processor and 16 GB RAM.

## Results

### Baseline clinical and biochemical characteristics

The present cohort consists of 37540 patients (37.5% males; median age of first blood pressure test: 63.7 years old, 95% CI: 50.5-73.0). The baseline clinical and biochemical parameters of the cohort are presented in **Table 1**. Within the present cohorts, hypertension is the most common comorbidity, followed by cardiovascular, respiratory, gastrointestinal diseases, and diabetes mellitus. In terms of drug prescription characteristics, 70.5% patients were prescribed with cardiovascular drugs while 21. 9% patients were treated with anti-diabetic medications. The use of both cardiovascular and anti-diabetic medications was more frequent in those who died than in those who did not, and in those who met the secondary outcomes of heart failure, acute myocardial infarction and TIA/stroke.

**Table 1.** Clinical and biochemical characteristics of patients with regard to mortality, HF, AMI and TIA/Stroke.

| **Variables** | **All patients (n=37540)**<br>**Median (IQR) or count**<br>**(%)** | **Mortality (n=15137)**<br>**Median (IQR) or count**<br>**(%)** | **HF (n=4086)**<br>**Median (IQR) or count**<br>**(%)** | **AMI (n=1333)**<br>**Median (IQR) or count**<br>**(%)** | **TIA/Stroke (n=6451)**<br>**Median (IQR) or count**<br>**(%)** | **P value** |
| --- | --- | --- | --- | --- | --- | --- |
| **Demographics** |  |  |  |  |  |  |
| Male sex | 14080(37.50%) | 7032(46.45%) | 1747(42.75%) | 544(40.81%) | 2828(43.8%) | 0.0102* |
| Age (year) | 63.7(50.5-73.0) | 73.(67.3-78.9) | 71.3(64.2-77.0) | 71.1(62.7-77.3) | 68.8(60.2-75.3) | 0.0311* |
| **Prior comorbidities** |  |  |  |  |  |  |
| Cardiovascular | 1713(4.56%) | 1222(8.07%) | 453(11.08%) | 123(9.22%) | 992(15.37%) | 0.0019** |
| Respiratory | 1859(4.95%) | 1370(9.05%) | 385(9.42%) | 85(6.37%) | 487(7.54%) | 0.0108* |
| Kidney | 330(0.87%) | 260(1.71%) | 72(1.76%) | 16(1.20%) | 93(1.44%) | 0.1240 |
| Endocrine | 74(0.19%) | 40(0.26%) | 12(0.29%) | 3(0.22%) | 16(0.24%) | 0.0231* |
| Diabetes mellitus | 331(0.88%) | 222(1.46%) | 56(1.37%) | 19(1.42%) | 92(1.42%) | 0.1196 |
| Hypertension | 2488(6.62%) | 1666(11.00%) | 463(11.33%) | 161(12.07%) | 809(12.54%) | 0.2581 |
| Gastrointestinal | 1602(4.26%) | 880(5.81%) | 238(5.82%) | 77(5.77%) | 364(5.64%) | 0.5612 |
| **Medications** |  |  |  |  |  |  |
| Cardiovascular drug use | 26462(70.49%) | 12622(83.38%) | 3452(84.48%) | 1110(83.27%) | 5772(89.47%) | 0.0481* |
| Diabetic mellitus drug use | 8218(21.89%) | 4387(28.98%) | 1144(27.99%) | 442(33.15%) | 1831(28.38%) | 0.0005*** |
| **Diastolic BP and its variability measures** |  |  |  |  |  |  |
| Baseline, mmHg | 74.0(66.0-81.2) | 73.0(65.0-80.2) | 73.0(66.0-81.0) | 72.0(66.0-80.3) | 73.0(66.0-80.1) | 0.1114 |
| Latest, mmHg | 72.0(65.0-80.0) | 71.0(64.0-80.0) | 71.0(64.0-80.1) | 72.0(64.0-80.0) | 72.0(65.0-80.2) | 0.5829 |
| Maximum, mmHg | 80.0(72.0-88.2) | 80.0(72.0-88.0) | 80.0(73.0-89.0) | 80.0(72.0-87.3) | 80.0(73.0-89.0) | 0.1571 |
| Minimal, mmHg | 67.0(60.0-74.0) | 65.0(58.0-72.0) | 65.0(58.0-72.0) | 65.0(58.0-72.0) | 65.0(59.0-72.2) | 0.7199 |
| Mean, mmHg | 73.3(67.0-80.0) | 72.5(66.4-78.9) | 72.77(66.58-79.3) | 72.5(66.0-78.6) | 73.0(66.8-79.3) | 0.3849 |
| Median, mmHg | 73.0(67.0-80.0) | 72.0(66.0-79.5) | 72.5(66.0-80.0) | 72.0(66.0-79.0) | 73.0(66.5-80.0) | 0.2795 |
| Variance | 40.9(19.6-75.7) | 49.2(23.6-87.6) | 49.67(23.3-87.2) | 50.0(23.3-85.0) | 45.0(22.3-82.6) | 0.3386 |
| SD | 6.4(4.4-8.7) | 7.0(4.9-9.4) | 7.1(4.8-9.3) | 7.1(4.8-9.2) | 6.7(4.7-9.1) | 0.0151* |
| RMS | 73.6(67.0-80.0) | 72.8(66.8-79.03) | 73.1(66.9-79.6) | 72.9(66.6-79.0) | 73.4(67.0-79.67) | 0.0183* |
| CV | 0.06(0.02-0.10) | 0.07(0.03-0.11) | 0.07(0.03-0.11) | 0.07(0.028-0.108) | 0.07(0.03-0.10) | 0.2510 |
| Variability Score, % | 50.0(33.3-66.7) | 52.1(33.1-66.8) | 50.1(32.1-64.4) | 51.2(33.8-63.2) | 49.8(31.1-65.5) | 0.0012** |
| **Systolic BP and its variability measures** |  |  |  |  |  |  |
| Baseline, mmHg | 135.0(120.0-150.0) | 141.0(128.0-155.0) | 140.0(128.0-155.0) | 140.0(126.0-153.0) | 139.0(124.0-152.0) | 0.1184 |
| Latest, mmHg | 134.0(120.0-148.0) | 140.0(126.0-152.0) | 140.0(125.0-152.0) | 139.0(125.0-151.0) | 138.0(123.5-150.0) | 0.0104* |
| Maximum, mmHg | 148.0(132.0-160.0) | 154.0(140.0-167.0) | 154.0(140.0-166.0) | 152.0(139.0-164.0) | 151.0(139.0-164.0) | 0.0092** |
| Minimal, mmHg | 122.0(110.0-135.0) | 127.0(114.0-140.0) | 126.0(113.0-139.0) | 124.0(113.0-137.0) | 123.0(112.0-136.0) | 0.0221* |
| Mean, mmHg | 135.5(123.25-146.3333) | 140.6(130.0-150.6667) | 140.2857(129.75-150.0) | 139.0(129.0-149.0) | 138.3333(128.0-148.2727) | 0.0107* |
| Median, mmHg | 135.0(122.0-146.5) | 140.0(130.0-150.5) | 140.0(130.0-150.0) | 139.5(128.0-150.0) | 138.5(128.0-149.0) | 0.0078** |
| Variance | 125.48(56.2-241.1) | 162.0(76.0-309.0) | 156.33(72.0-296.1) | 146.03(72.0-278.0) | 144.5(70.1-267.0) | 0.4186 |
| SD | 11.20(7.5-15.5) | 12.7(8.7-17.6) | 12.5(8.5-17.21) | 12.1(8.5-16.7) | 12.02(8.4-16.3) | 0.4279 |
| RMS | 136.0(123.7-146.9) | 141.2(130.5-151.2) | 140.8(130.0-150.8) | 139.5(129.6-149.4) | 138.9(128.4-148.9) | 0.0203* |
| CV | 0.06(0.02-0.09) | 0.07(0.03-0.10) | 0.07(0.03-0.10) | 0.07(0.03-0.10) | 0.07(0.03-0.10) | 0.2273 |
| Variability Score, % | 60.0(50.0-75.1) | 66.7(52.0-76.3) | 62.7(51.0-73.3) | 63.5(52.2-75.2) | 64.3(50.5-73.3) | 0.0311* |
| **CBC tests** |  |  |  |  |  |  |
| MCV, fL | 90.0(86.5-93.2); n=10106 | 90.7(87.0-94.0); n=5069 | 90.4(86.9-93.6); n=1276 | 90.4(86.85-93.35); n=455 | 90.4(87.0-93.5); n=2198 | 0.2102 |
| Basophil, x10<sup>9</sup>/L | 0.02(0.0-0.4); n=4326 | 0.021(0.0-0.042); n=2498 | 0.01(0.0-0.23); n=623 | 0.01(0.0-0.025); n=215 | 0.01(0.0-0.22); n=1040 | 0.0263* |
| Eosinophil, x10<sup>9</sup>/L | 0.1(0.1-0.2); n=5313 | 0.1(0.1-0.2); n=2939 | 0.1(0.1-0.2); n=733 | 0.1(0.1-0.2); n=248 | 0.1(0.1-0.2); n=1209 | 0.9145 |
| Lymphocyte, x10<sup>9</sup>/L | 1.7(1.3-2.2); n=5351 | 1.6(1.1-2.1); n=2966 | 1.6(1.1-2.1); n=737 | 1.8(1.4-2.3); n=251 | 1.7(1.2-2.2); n=1213 | 0.0007*** |
| Blast, x10<sup>9</sup>/L | 0.0(0.0-0.08); n=971 | 0.0(0.0-0.1); n=441 | 0.0(0.0-0.1); n=111 | 0.0(0.0-0.1); n=34 | 0.0(0.0-0.06); n=171 | 0.1588 |
| Metamyelocyte, x10<sup>9</sup>/L | 0.16(0.09-0.5); n=22 | 0.17(0.09-0.4); n=17 | 0.15(0.08-0.42); n=4 | 1.6(1.6-1.6); n=1 | 0.17(0.08-0.44); n=9 | 0.0019** |
| Monocyte, x10<sup>9</sup>/L | 0.5(0.4-0.6); n=5349 | 0.5(0.4-0.7); n=2965 | 0.5(0.4-0.7); n=737 | 0.5(0.4-0.6); n=251 | 0.5(0.4-0.7); n=1213 | 0.5711 |
| Neutrophil, x10<sup>9</sup>/L | 4.5(3.4-6.2); n=5344 | 4.9(3.7-7.1); n=2964 | 4.8(3.7-6.8); n=737 | 4.5(3.6-6.4); n=251 | 4.7(3.6-6.6); n=1213 | 0.0188* |
| WBC, x10<sup>9</sup>/L | 7.2(5.9-8.86); n=10268 | 7.5(6.1-9.5); n=5167 | 7.485(6.155-9.3); n=1308 | 7.5(6.2-9.5); n=465 | 7.4(6.1-9.2); n=2230 | 0.3034 |
| MCH, pg | 30.5(29.2-31.7); n=10105 | 30.7(29.3-31.9); n=5069 | 30.6(29.2-31.8); n=1276 | 30.5(29.3-31.7); n=455 | 30.6(29.4-31.8); n=2197 | 0.8309 |
| Myelocyte, x10<sup>9</sup>/L | 0.17(0.08-0.3); n=26 | 0.17(0.1-0.3); n=22 | 1.31(1.31-1.31); n=1 | 0.16(0.16-0.16); n=1 | 0.15(0.07-0.17); n=10 | 0.0002*** |
| Platelet, x10<sup>9</sup>/L | 229.0(189.0-274.0); n=10268 | 220.0(178.0-268.0); n=5167 | 219.0(180.0-262.0); n=1308 | 224.0(179.0-268.0); n=465 | 220.0(181.0-265.5); n=2230 | 0.6518 |
| Reticulocyte, x10<sup>9</sup>/L | 50.7(35.19-77.7); n=207 | 50.1(33.3-78.2); n=124 | 44.195(35.3-64.98); n=18 | 65.1(43.0-70.7); n=13 | 50.08(33.4-70.6); n=46 | <0.0001**<br>* |
| RBC, x10<sup>12</sup>/L | 4.35(4.0-4.72); n=10092 | 4.23(3.84-4.63); n=5065 | 4.29(3.92-4.68); n=1273 | 4.3(3.94-4.65); n=454 | 4.33(3.95-4.69); n=2194 | 0.6980 |
| HCT, L/L | 0.39(0.36-0.41); n=8924 | 0.38(0.35-0.41); n=4478 | 0.38(0.35-0.41); n=1149 | 0.38(0.35-0.41); n=418 | 0.39(0.36-0.42); n=1932 | 0.0444* |
| **Liver and renal function tests** |  |  |  |  |  |  |
| K/Potassium, mmol/L | 4.2(3.9-4.5); n=15515 | 4.2(3.8-4.5); n=7189 | 4.2(3.8-4.5); n=1906 | 4.2(3.8-4.5); n=632 | 4.2(3.9-4.5); n=3227 | 0.4302 |
| Urate, mmol/L | 0.36(0.29-0.45); n=3083 | 0.39(0.31-0.48); n=1482 | 0.4(0.33-0.495); n=430 | 0.39(0.3139-0.46); n=131 | 0.38(0.31-0.47); n=659 | 0.0092** |
| Albumin, g/L | 41.0(38.6-44.0); n=13361 | 40.0(37.0-43.0); n=6265 | 40.0(37.5-43.0); n=1625 | 41.0(38.0-44.0); n=547 | 41.0(38.0-43.0); n=2790 | 0.1140 |
| Na/Sodium, mmol/L | 141.0(139.0-142.0); n=15593 | 141.0(138.0-142.0); n=7225 | 141.0(139.0-142.0); n=1917 | 141.0(139.0-143.0); n=635 | 141.0(139.0-142.0); n=3243 | 0.3870 |
| Urea, mmol/L | 5.6(4.6-7.0); n=15592 | 6.3(5.1-8.0); n=7224 | 6.2(5.1-7.8); n=1916 | 6.2(4.8-7.9); n=636 | 6.0(4.9-7.5); n=3243 | 0.6540 |
| Protein, g/L | 74.0(71.0-78.0); n=13220 | 73.0(69.0-77.0); n=6212 | 73.0(70.0-77.0); n=1617 | 74.0(70.0-77.8); n=540 | 73.25(70.0-77.0); n=2768 | 0.1558 |
| Creatinine, µmol/L | 85.0(73.0-102.0); n=15592 | 95.0(80.0-115.0); n=7224 | 93.0(78.0-113.0); n=1916 | 92.0(78.0-113.0); n=636 | 90.0(77.0-108.0); n=3243 | 0.0003*** |
| ALP, U/L | 79.0(65.0-98.0); n=13085 | 84.0(69.0-104.0); n=6153 | 81.0(67.0-99.0); n=1597 | 83.0(66.0-101.0); n=537 | 81.0(67.0-99.0); n=2749 | 0.1519 |
| Aspartate Transaminase, U/L | 22.0(18.0-29.0); n=2191 | 23.0(18.0-30.0); n=1113 | 22.0(18.0-29.0); n=287 | 22.0(17.0-30.0); n=93 | 22.0(18.0-29.0); n=531 | 0.2090 |
| ALT, U/L | 20.0(14.0-29.0); n=11494 | 18.0(13.0-27.0); n=5424 | 19.0(14.0-27.0); n=1438 | 18.0(13.0-27.0); n=500 | 19.0(14.0-28.0); n=2424 | 0.0173* |
| Bilirubin, µmol/L | 9.3(7.0-12.8); n=13196 | 9.0(7.0-13.0); n=6207 | 9.4(7.0-12.7); n=1612 | 8.9(6.6-12.7); n=541 | 9.4(7.0-12.85); n=2768 | 0.2399 |
| **Diabetes and lipid tests** |  |  |  |  |  |  |
| HbA1c, % | 12.8(11.3-13.9); n=10308 | 12.5(10.9-13.7); n=5182 | 12.7(11.2-13.9); n=1314 | 12.8(11.4-13.9); n=466 | 12.8(11.15-13.9); n=2235 | 0.0101* |
| Cholesterol, mmol/L | 5.3(4.65-5.96); n=10579 | 5.2(4.56-5.8964); n=4323 | 5.26(4.565-5.9); n=1215 | 5.39(4.61-6.0); n=403 | 5.3(4.6-5.92); n=2335 | 0.6443 |
| Glucose, mmol/L | 6.0(5.2-7.7); n=13886 | 6.4(5.4-8.4); n=6382 | 6.3(5.3-8.35); n=1712 | 6.4(5.4-8.2); n=570 | 6.2(5.3-8.0); n=2999 | 0.2806 |
| Triglycerides, mmol/L | 1.45(1.02-2.09); n=10493 | 1.43(1.0145-2.05); n=4276 | 1.44(1.0264-2.08); n=1206 | 1.525(1.02-2.045); n=400 | 1.5(1.08-2.13); n=2315 | 0.0387* |

### Blood Pressure Variability Measures

At least three blood pressure measurements were available in 29597 patients, and the median number of measurements per patient was 5 (95% CI: [3,7]). Overall, baseline diastolic blood pressure was 74.0 [66.0-81.0] mmHg. In terms of its variability measures, they are represented by 1) patient-specific mean 73.3 [67.0-80.0] mmHg; 2) median 73.0 [67.0-80.0] mmHg; 3) variance 40.9 [19.6-75.7]; 4) SD 6.4 [4.4-8.7]; 5) RMS 73.6 [67.0-80.0]; 6) CV 0.06 [0.02-0.10]; and 7) variability score (%) 50.0 [33.3-66.7]. Overall, baseline systolic blood pressure was 135.0 [120.0-150.0] mmHg. In terms of its variability measures, they are represented by 1) patient-specific mean 135.5 [123.3-146.3] mmHg; 2) median 135.0 [122.0-146.5] mmHg; 3) variance 125.5 [56.2-241.1]; 4) SD 11.2 [7.5-15.5]; 5) RMS 136.0 [123.7-146.9]; 6) CV 0.06 [0.02-0.09]; and 7) variability score (%) 60.0 [50.0-75.1]. Further details on blood pressure and its variability measures are provided in **Table 1**.

### Outcomes and Cox regression

On follow-up, a total of 15137 deaths were recorded. For secondary outcomes, 4086 had incident heart failure, 1333 had acute myocardial infarction and 6451 had TIA/stroke. The Kaplan-Meier survival curves of primary mortality and secondary complication outcomes are shown in **Figure 1**. Males have a shorter time-to-event for all primary and secondary outcomes. The univariate Cox regression analysis for predicting primary mortality and secondary complication outcomes are presented in **Table 2**.

**Table 2.** Identified significant predictors through univariate Cox regression analysis. * for p≤ 0.05, ** for p ≤ 0.01, *** for p ≤ 0.001

| Variable | Mortality<br>HR (95% CI) | P value | HF<br>HR (95% CI) | P value | AMI<br>HR (95% CI) | P value | TIA/Stroke<br>HR (95% CI) | P value |
| --- | --- | --- | --- | --- | --- | --- | --- | --- |
| **Demographics** |  |  |  |  |  |  |  |  |
| Male sex | 1.65[1.60,1.71] | <0.0001*** | 1.26[1.19,1.35] | <0.0001*** | 1.15[1.03,1.29] | 0.0107* | 1.34[1.28,1.41] | <0.0001*** |
| Age (year) | 1.10[1.10,1.11] | <0.0001*** | 1.05[1.04,1.06] | <0.0001*** | 1.04[1.04,1.05] | <0.0001*** | 1.03[1.03,1.04] | <0.0001*** |
| **Prior comorbidities** |  |  |  |  |  |  |  |  |
| Cardiovascular | 1.017[1.01,1.018] | <0.0001*** | 3.075[2.79,3.39] | <0.0001*** | 2.20[1.83,2.65] | <0.0001*** | 6.36[5.94,6.80] | <0.0001*** |
| Respiratory | 1.02[1.02,1.021] | <0.0001*** | 2.22[2.00,2.47] | <0.0001*** | 1.32[1.06,1.65] | 0.0124* | 1.78[1.62,1.95] | <0.0001*** |
| Kidney | 1.022[1.02,1.023] | <0.0001*** | 2.29[1.81,2.89] | <0.0001*** | 1.40[0.85,2.29] | 0.185 | 1.95[1.59,2.39] | <0.0001*** |
| Endocrine | 1.014[1.01,1.02] | <0.0001*** | 1.63[0.93,2.87] | 0.0909 | 1.15[0.37,3.58] | 0.806 | 1.34[0.82,2.19] | 0.239 |
| Diabetes mellitus | 1.016[1.014,1.02] | <0.0001*** | 1.66[1.27,2.16] | 0.0002*** | 1.66[1.06,2.62] | 0.0277* | 1.83[1.49,2.25] | <0.0001*** |
| Hypertension | 1.016[1.015,1.02] | <0.0001*** | 1.93[1.75,2.13] | <0.0001*** | 1.99[1.68,2.34] | <0.0001*** | 2.41[2.24,2.59] | <0.0001*** |
| Gastrointestinal | 1.011[1.01,1.012] | <0.0001*** | 1.44[1.27,1.64] | <0.0001*** | 1.39[1.10,1.75] | 0.00513** | 1.45[1.30,1.61] | <0.0001*** |
| **Medications** |  |  |  |  |  |  |  |  |
| Cardiovascular drug use | 2.51[2.41,2.62] | <0.0001*** | 2.369[2.18,2.58] | <0.0001*** | 2.11[1.83,2.44] | <0.0001*** | 3.87[3.57,4.19] | <0.0001*** |
| Diabetes mellitus drug use | 1.62[1.57,1.68] | <0.0001*** | 1.411[1.32,1.51] | <0.0001*** | 1.79[1.60,2.01] | <0.0001*** | 1.45[1.38,1.53] | <0.0001*** |
| **Diastolic BP and its variability measures** |  |  |  |  |  |  |  |  |
| Baseline, mmHg | 0.99[0.99,0.995] | <0.0001*** | 0.999[0.996,1.00] | 0.514 | 0.994[0.99,0.999] | 0.00978** | 0.997[0.995,1.00] | 0.0028** |
| Latest, mmHg | 0.99[0.987,0.991] | <0.0001*** | 0.995[0.992,0.997] | 0.000152*** | 0.993[0.988,0.997] | 0.0027** | 0.997[0.995,1.00] | 0.0107* |
| Maximum, mmHg | 0.9998[0.9984,1.001] | 0.772 | 1.004[1.002,1.007] | 0.00187** | 0.999[0.994,1.00] | 0.5300 | 1.004[1.002,1.006] | 0.0002*** |
| Minimal, mmHg | 0.9839[0.9824,0.9854] | <0.0001*** | 0.989[0.986,0.99] | <0.0001*** | 0.986[0.981,0.991] | <0.0001*** | 0.99[0.988,0.99] | <0.0001*** |
| Mean, mmHg | 0.9892[0.9876,0.9909] | <0.0001*** | 0.996[0.993,1.00] | 0.0008*** | 0.991[0.985,0.996] | 0.00013*** | 0.997[0.994,1.00] | 0.0001*** |
| Median, mmHg | 0.9891[0.9875,0.9907] | <0.0001*** | 0.995[0.99,1.00] | 0.00071*** | 0.991[0.986,0.997] | 0.00014*** | 0.997[0.994,0.999] | 0.0003*** |
| Variance | 1.002[1.002,1.002] | <0.0001*** | 1.002[1.001,1.002] | <0.0001*** | 1.001[1.00,1.002] | 0.00342** | 1.001[1.001,1.001] | <0.0001*** |
| SD | 1.058[1.053,1.063] | <0.0001*** | 1.04[1.03,1.05] | <0.0001*** | 1.03[1.02,1.05] | <0.0001*** | 1.02[1.02,1.03] | <0.0001*** |
| RMS | 0.9899[0.9883,0.9916] | <0.0001*** | 0.996[0.993,1.00] | 0.0231* | 0.99[0.986,0.997] | 0.00226** | 0.997[0.995,1.00] | 0.035* |
| CV | 52.36[38.15,71.86] | <0.0001*** | 30.34[16.8,54.79] | <0.0001*** | 20.00[7.10,56.32] | <0.0001*** | 19.22[11.96,30.87] | <0.0001*** |
| Variability Score, % | 1.005[1.004,1.006] | <0.0001*** | 1.003[1.002,1.005] | <0.0001*** | 1.005[1.002,1.008] | 0.000226*** | 1.004[1.002,1.005] | <0.0001*** |
| **Systolic BP and its variability measures** |  |  |  |  |  |  |  |  |
| Baseline, mmHg | 1.017[1.016,1.017] | <0.0001*** | 1.013[1.012,1.014] | <0.0001*** | 1.009[1.007,1.012] | <0.0001*** | 1.007[1.006,1.009] | <0.0001*** |
| Latest, mmHg | 1.017[1.016,1.017] | <0.0001*** | 1.012[1.011,1.014] | <0.0001*** | 1.01[1.007,1.012] | <0.0001*** | 1.009[1.008,1.01] | <0.0001*** |
| Maximum, mmHg | 1.018[1.018,1.019] | <0.0001*** | 1.014[1.013,1.016] | <0.0001*** | 1.01[1.008,1.013] | <0.0001*** | 1.01[1.009,1.011] | <0.0001*** |
| Minimal, mmHg | 1.016[1.015,1.017] | <0.0001*** | 1.011[1.009,1.012] | <0.0001*** | 1.008[1.005,1.01] | <0.0001*** | 1.006[1.004,1.007] | <0.0001*** |
| Mean, mmHg | 1.024[1.023,1.025] | <0.0001*** | 1.018[1.016,1.02] | <0.0001*** | 1.013[1.01,1.016] | <0.0001*** | 1.012[1.01,1.013] | <0.0001*** |
| Median, mmHg | 1.023[1.023,1.024] | <0.0001*** | 1.017[1.016,1.019] | <0.0001*** | 1.013[1.01,1.016] | <0.0001*** | 1.012[1.01,1.013] | <0.0001*** |
| Variance | 1.001[1.001,1.001] | <0.0001*** | 1.001[1,1.001] | <0.0001*** | 1[1,1.001] | <0.0001*** | 1[1,1] | <0.0001*** |
| SD | 1.043[1.041,1.045] | <0.0001*** | 1.028[1.024,1.033] | <0.0001*** | 1.022[1.014,1.03] | <0.0001*** | 1.019[1.016,1.023] | <0.0001*** |
| RMS | 1.024[1.024,1.025] | <0.0001*** | 1.018[1.016,1.02] | <0.0001*** | 1.013[1.01,1.016] | <0.0001*** | 1.012[1.011,1.013] | <0.0001*** |
| CV | 72.06[51.75,100.3] | <0.0001*** | 31.9[17.2,59.4] | <0.0001*** | 15.84[5.335,47.01] | <0.0001*** | 28.62[17.45,46.94] | <0.0001*** |
| Variability Score, % | 1.007[1.006,1.008] | <0.0001*** | 1.005[1.003,1.007] | <0.0001*** | 1.006[1.003,1.009] | 0.000279*** | 1.006[1.005,1.008] | <0.0001*** |
| **Complete blood counts** |  |  |  |  |  |  |  |  |
| MCV, fL | 1.019[1.015,1.023] | <0.0001*** | 1.005[0.998,1.01] | 0.144 | 1.011[0.9988,1.023] | 0.0781. | 1.011[1.006,1.017] | <0.0001*** |
| Basophil, x10<sup>9</sup>/L | 1.344[0.5361,3.371] | 0.528 | 0.27[0.04,1.9] | 0.19 | 0.41[0.016,10.69] | 0.593 | 1.032[0.2522,4.225] | 0.965 |
| Eosinophil, x10<sup>9</sup>/L | 1.17[1.04,1.31] | 0.00781** | 1.35[1.15,1.58] | 0.0003*** | 1.13[0.75,1.70] | 0.569 | 1.06[0.86,1.31] | 0.603 |
| Lymphocyte, x10<sup>9</sup>/L | 0.6261[0.5928,0.6614] | <0.0001*** | 0.838[0.76,0.93] | 0.000671*** | 1.019[0.9737,1.067] | 0.411 | 0.9826[0.9251,1.044] | 0.568 |
| Blast, x10<sup>9</sup>/L | 16[2.254,113.5] | 0.00555** | 6.06[0.14,263.5] | 0.349 | 1313[5.274,326912] | 0.0108* | 0.6215[0.02431,15.89] | 0.774 |
| Metamyelocyte, x10<sup>9</sup>/L | 1.102[0.4132,2.941] | 0.846 | 0.36[0.02,6.06] | 0.481 | 38.42[0.09013,16373] | 0.238 | 0.8961[0.2493,3.221] | 0.867 |
| Monocyte, x10<sup>9</sup>/L | 2.305[2.086,2.546] | <0.0001*** | 1.72[1.4,2.11] | <0.0001*** | 1.332[0.889,1.995] | 0.165 | 1.549[1.307,1.836] | <0.0001*** |
| Neutrophil, x10<sup>9</sup>/L | 1.096[1.087,1.105] | <0.0001*** | 1.04[1.02,1.06] | <0.0001*** | 1.005[0.9675,1.044] | 0.797 | 1.034[1.018,1.051] | <0.0001*** |
| WBC, x10<sup>9</sup>/L | 1.054[1.049,1.059] | <0.0001*** | 1.032[1.02,1.05] | <0.0001*** | 1.026[1.004,1.048] | 0.022* | 1.026[1.016,1.036] | <0.0001*** |
| MCH, pg | 1.027[1.017,1.037] | <0.0001*** | 1.004[0.987,1.02] | 0.623 | 1.022[0.9909,1.053] | 0.17 | 1.026[1.012,1.041] | 0.000257*** |
| Myelocyte, x10<sup>9</sup>/L | 2.487[0.9267,6.677] | 0.0705. | 2.88[0.59,13.94] | 0.19 | 0.1058[4.15E-07,26974] | 0.724 | 0.01254[7.35E-05,2.141] | 0.095. |
| Platelet, x10<sup>9</sup>/L | 0.9975[0.9971,0.9979] | <0.0001*** | 0.998[0.998,0.999] | <0.0001*** | 0.9988[0.9975,1] | 0.0807. | 0.9981[0.9975,0.9987] | <0.0001*** |
| Reticulocyte, x10<sup>9</sup>/L | 0.9981[0.9928,1.003] | 0.49 | 0.998[0.984,1.01] | 0.735 | 1.001[0.9855,1.017] | 0.91 | 0.9954[0.9864,1.005] | 0.325 |
| RBC, x10<sup>12</sup>/L | 0.5392[0.5139,0.5658] | <0.0001*** | 0.819[0.749,0.895] | <0.0001*** | 0.8485[0.7327,0.9826] | 0.0282* | 0.8823[0.8251,0.9436] | 0.000256*** |
| HCT, L/L | 0.0028[0.0015,0.0051] | <0.0001*** | 0.116[0.0366,0.367] | 0.000248*** | 0.3408[0.0501,2.318] | 0.271 | 0.4982[0.2013,1.233] | 0.132 |
| **Liver and renal function tests** |  |  |  |  |  |  |  |  |
| K/Potassium, mmol/L | 0.86[0.82,0.90] | <0.0001*** | 0.881[0.81,0.959] | 0.00355** | 0.917[0.7919,1.062] | 0.247 | 0.9398[0.8806,1.003] | 0.0616. |
| Urate, mmol/L | 18.1[11.73,27.93] | <0.0001*** | 25.64[12.06,54.49] | <0.0001*** | 3.887[0.9445,16] | 0.06. | 5.874[3.117,11.07] | <0.0001*** |
| Albumin, g/L | 0.88[0.88,0.89] | <0.0001*** | 0.957[0.947,0.967] | <0.0001*** | 0.9801[0.9619,0.9987] | 0.0362* | 0.9691[0.9612,0.9771] | <0.0001*** |
| Na/Sodium, mmol/L | 0.94[0.94,0.95] | <0.0001*** | 0.987[0.973,0.99] | 0.0484* | 1.008[0.9833,1.033] | 0.536 | 0.9941[0.9836,1.005] | 0.281 |
| Urea, mmol/L | 1.118[1.114,1.123] | <0.0001*** | 1.058[1.048,1.068] | <0.0001*** | 1.055[1.036,1.073] | <0.0001*** | 1.054[1.045,1.063] | <0.0001*** |
| Protein, g/L | 0.94[0.94,0.95] | <0.0001*** | 0.974[0.966,0.982] | <0.0001*** | 0.9814[0.9673,0.9956] | 0.0107* | 0.9757[0.9695,0.982] | <0.0001*** |
| Creatinine, µmol/L | 1.005[1.005,1.005] | <0.0001*** | 1.003[1.002,1.003] | <0.0001*** | 1.002[1.001,1.003] | <0.0001*** | 1.003[1.003,1.004] | <0.0001*** |
| ALP, U/L | 1.004[1.004,1.004] | <0.0001*** | 1.001[1,1.002] | 0.00658** | 1.001[0.9999,1.002] | 0.0688. | 1[0.9997,1.001] | 0.228 |
| Aspartate Transaminase, U/L | 1[1,1.001] | 0.0291* | 1.001[0.99,1.002] | 0.0846. | 1.001[0.99,1.002] | 0.376 | 0.9999[0.9987,1.001] | 0.795 |
| ALT, U/L | 0.997[0.996,0.998] | <0.0001*** | 0.997[0.995,0.9996] | 0.0231* | 1.00[0.99,1.002] | 0.63 | 1.00[0.997,1.00] | 0.14 |
| Bilirubin, µmol/L | 1.007[1.005,1.01] | <0.0001*** | 0.998[0.992,1.005] | 0.609 | 0.99[0.971,1.001] | 0.0739. | 0.997[0.99,1.002] | 0.241 |
| **Diabetes and lipid tests** |  |  |  |  |  |  |  |  |
| HbA1c, % | 0.94[0.92,0.96] | <0.0001*** | 0.96[0.91,1.01] | 0.109 | 1.01[0.92,110] | 0.888 | 0.97[0.94,1.01] | 0.125 |
| Cholesterol, mmol/L | 0.98[0.98,0.99] | <0.0001*** | 1.005[0.99,1.018] | 0.429 | 1.03[1.003,1.05] | 0.0252* | 1.00[0.99,1.01] | 0.526 |
| Glucose, mmol/L | 1.06[1.06,1.07] | <0.0001*** | 1.05[1.03,1.06] | <0.0001*** | 1.03[1.01,1.05] | 0.00973** | 1.02[1.01,1.03] | <0.0001*** |
| Triglycerides, mmol/L | 0.986[0.963,1.01] | 0.24 | 1.00[0.96,1.05] | 0.845 | 0.98[0.91,1.06] | 0.621 | 1.02[0.995,1.05] | 0.114 |

**Figure 1.**
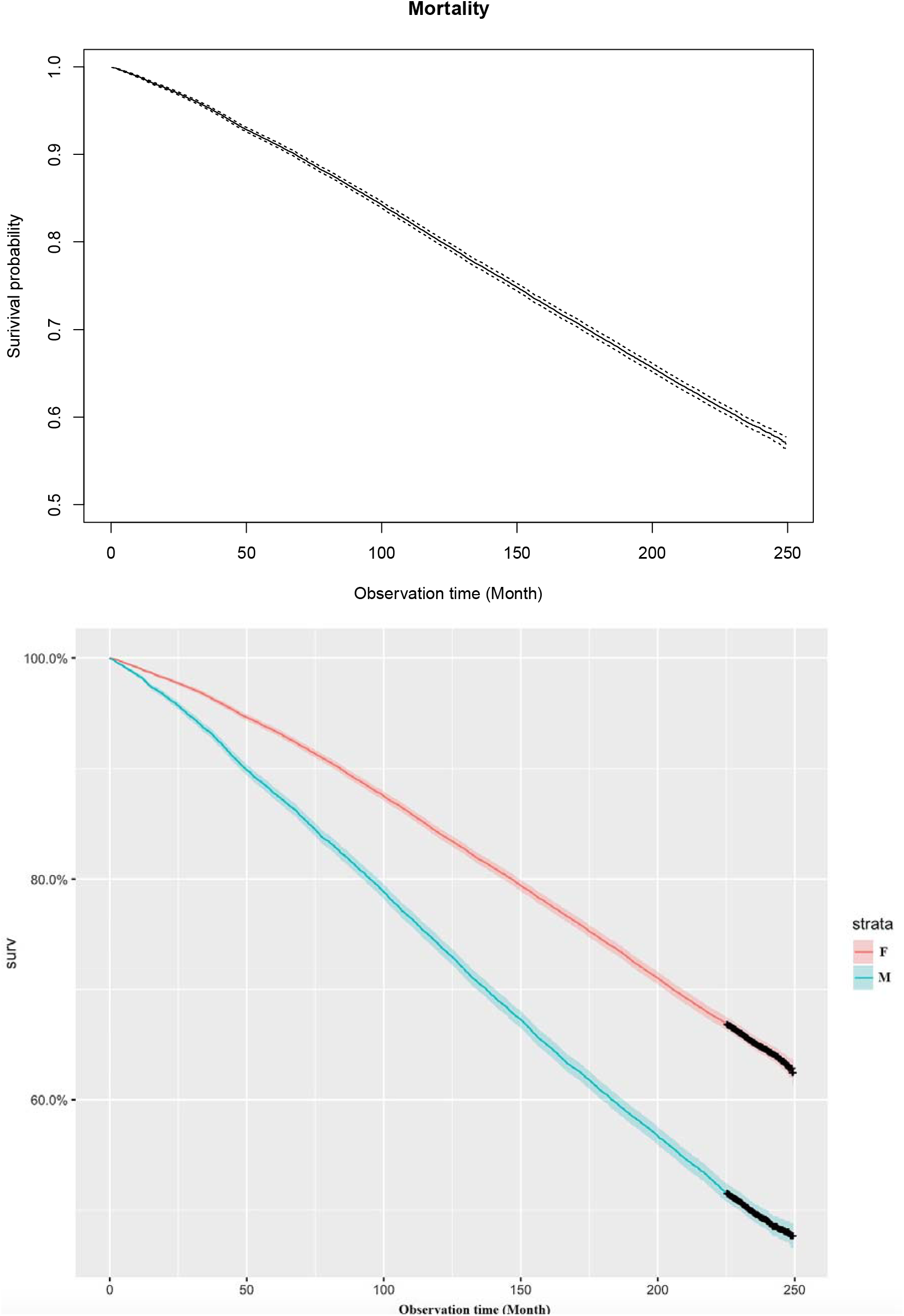
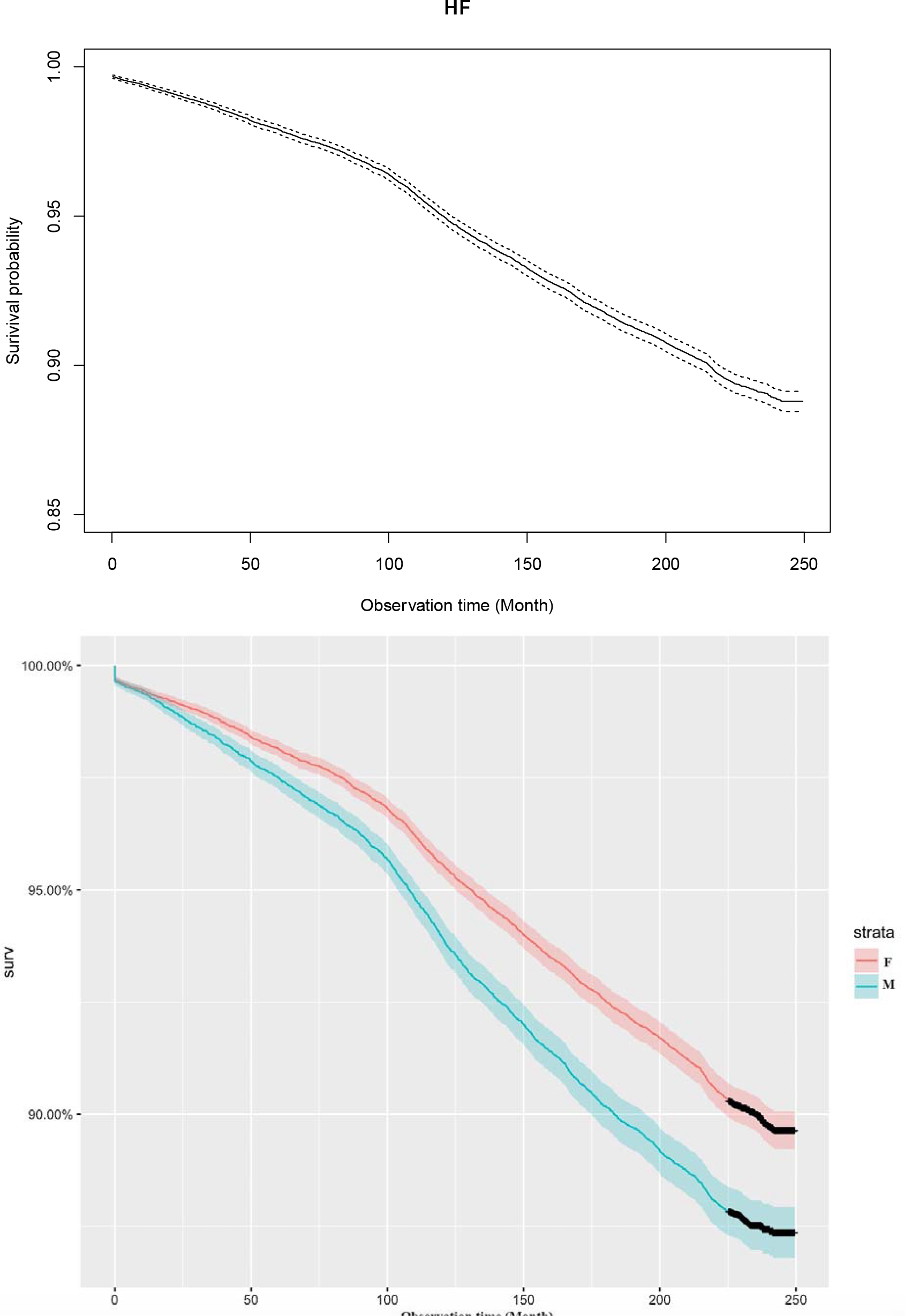
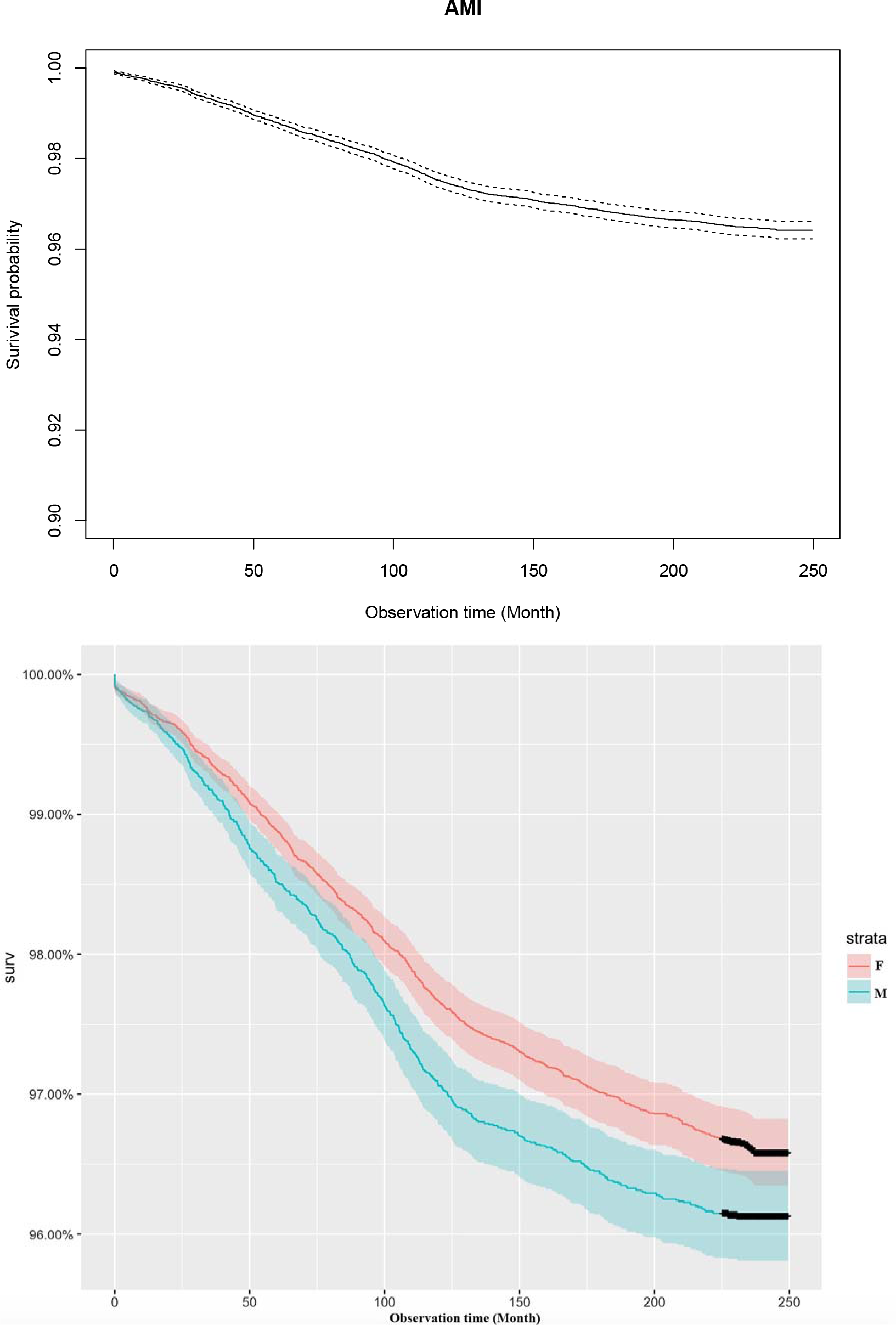
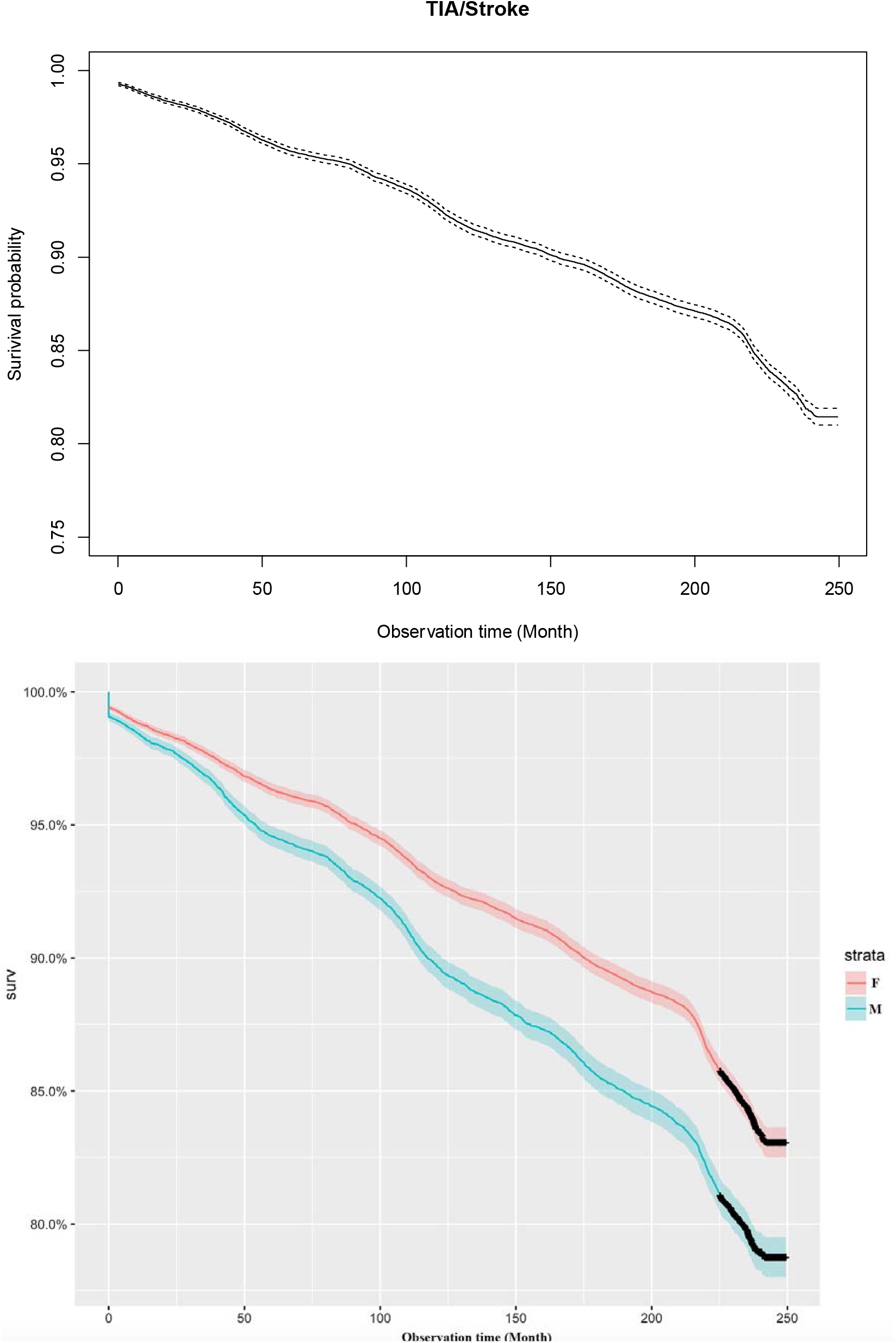
Kaplan-Meier survival curves of outcomes.

**Figure 2.**
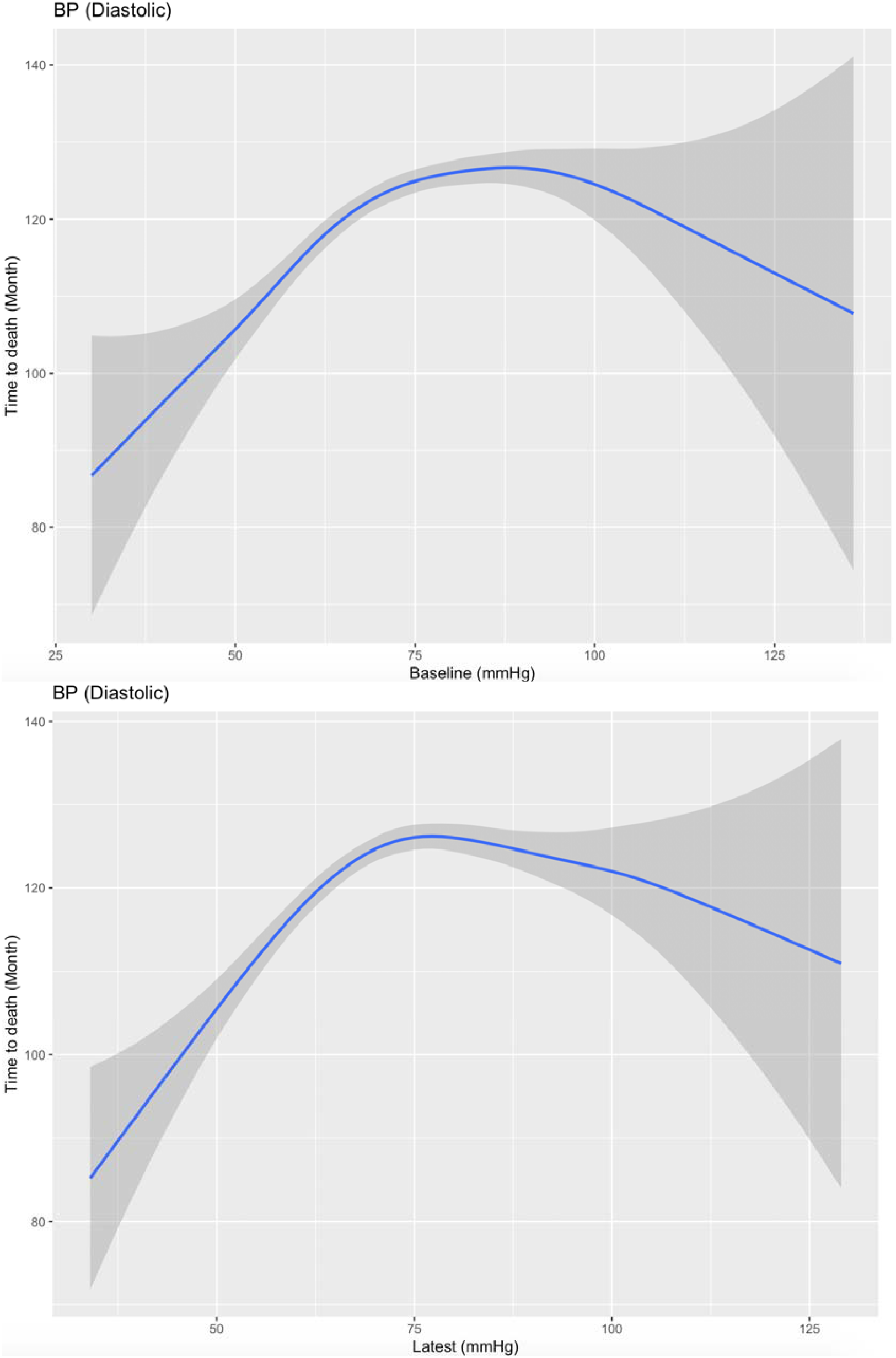
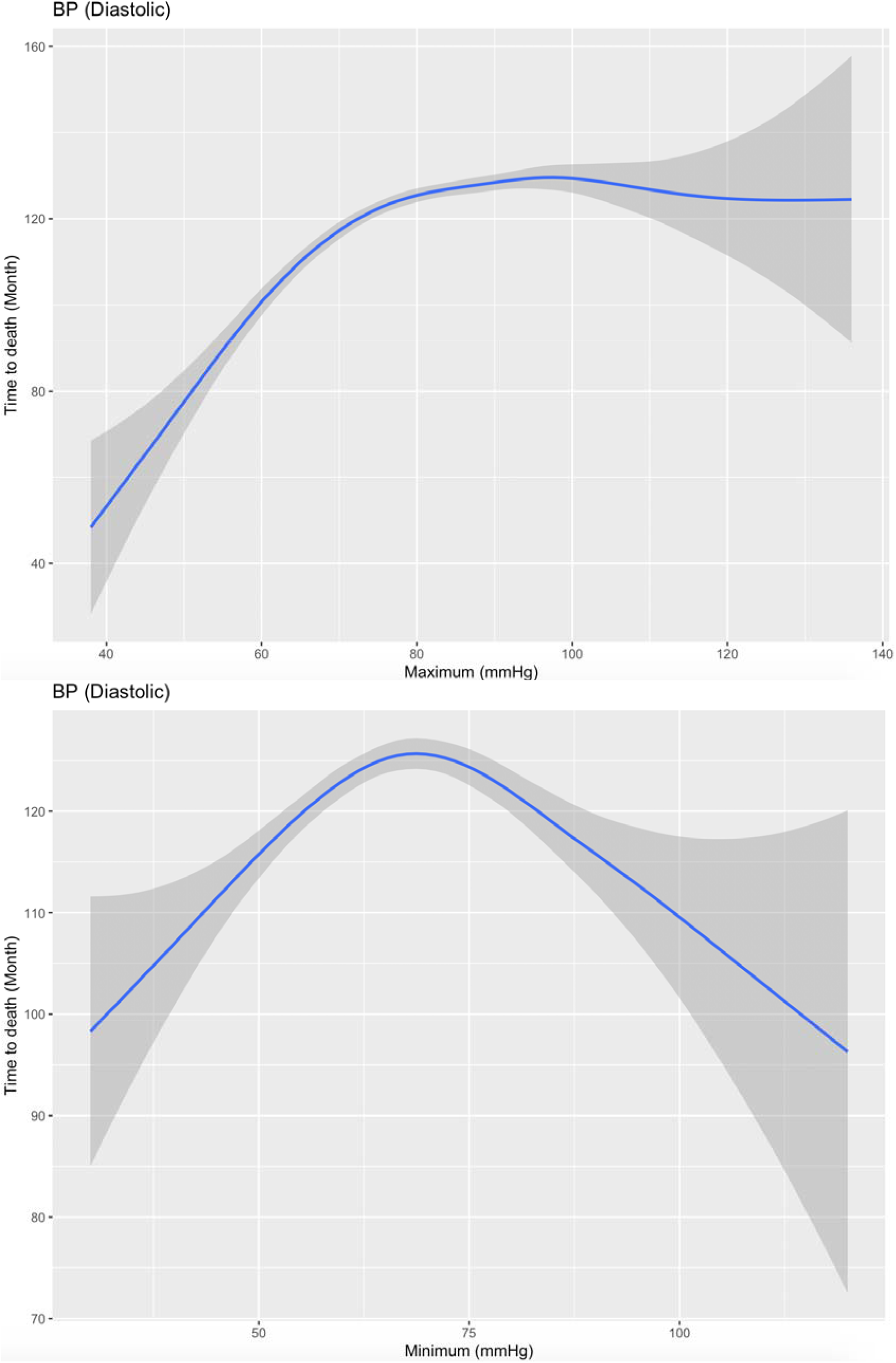
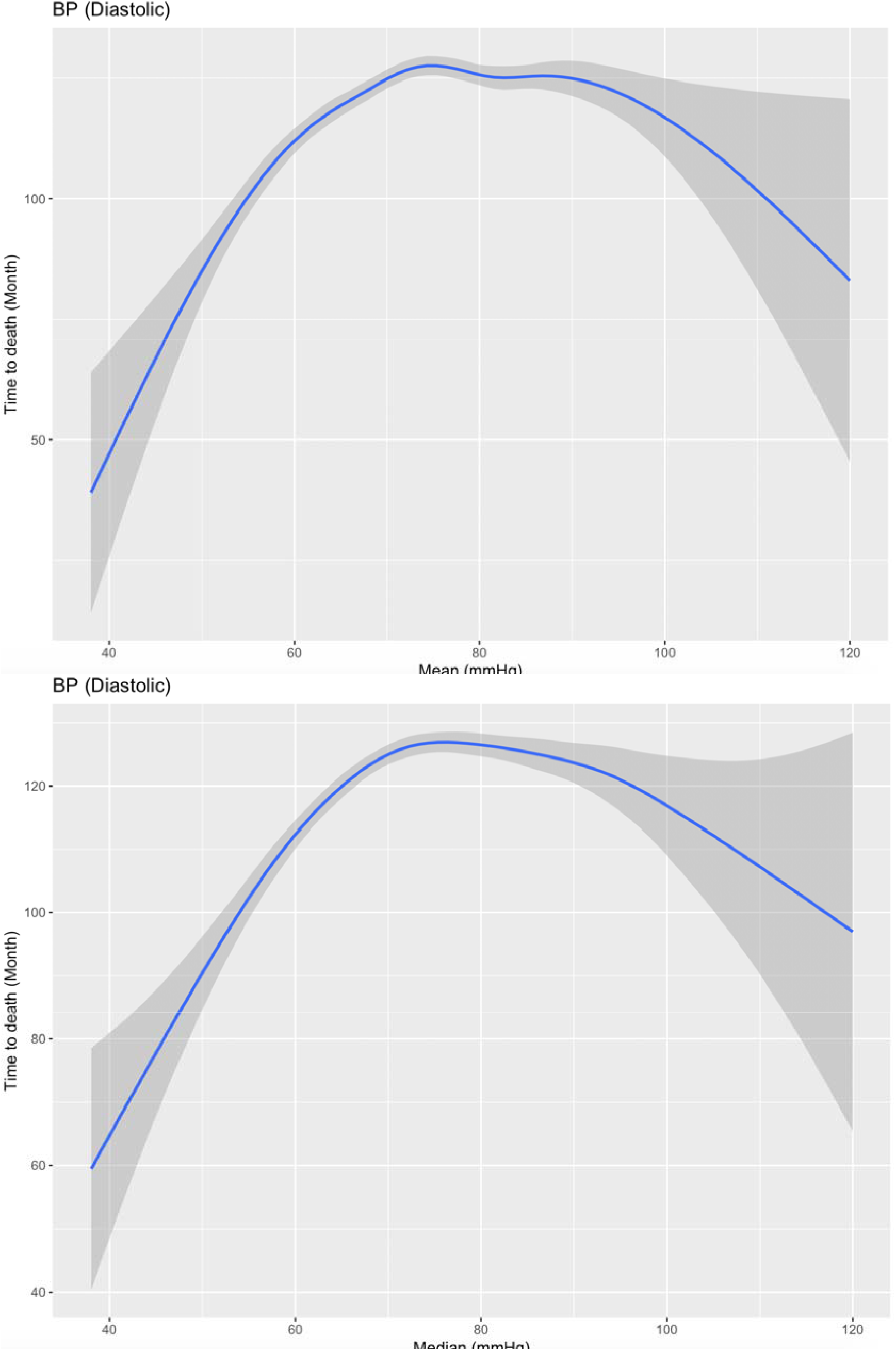
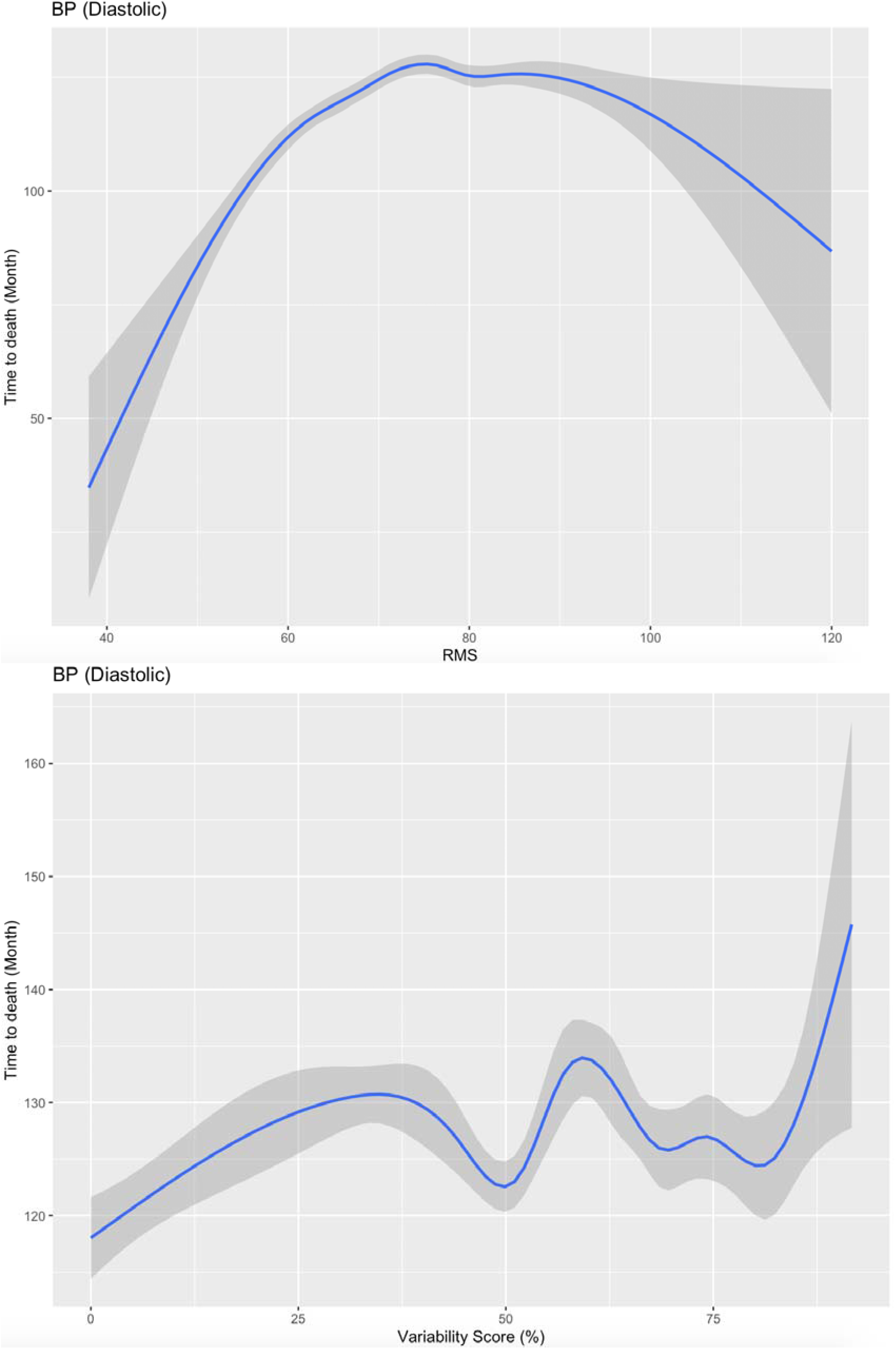
Associations of time-to-death of all-cause mortality with diastolic BP and its variability measures.

For predicting the primary outcome of all-cause mortality, diastolic blood pressure variance, SD, CV and variability score were found to be significant positive predictors, while all systolic blood pressure measures were all predictive (HR>1 and p value<0.001). In addition, diastolic blood pressure variance, SD, CV and variability score are significant positive predictors of incident heart failure, acute myocardial infarction and TIA/stroke. All systolic blood pressure measures including variability measures were significant positive predictors of incident heart failure, acute myocardial infarction and TIA/stroke.

The individual effects of blood pressure, in addition to blood pressure variability, on all-cause mortality was further examined. Results from a generalized additive model showed prolonged time-to-death at the extremes of the baseline, latest, mean and median diastolic blood pressure (**Figures 3**). The low baseline, latest, mean and median diastolic blood pressure subgroup had significantly shorter time-till-death for all-cause mortality (P < 0.001). On the contrary, a nonlinear M-shaped relationship between time-to-death and systolic blood pressure was observed: low baseline, latest, mean and median diastolic blood pressure were found in patients with both longer and shorter time-to-death duration, while patients in the middle had high examination results of baseline, latest, mean and median diastolic blood pressure.

**Figure 3.**
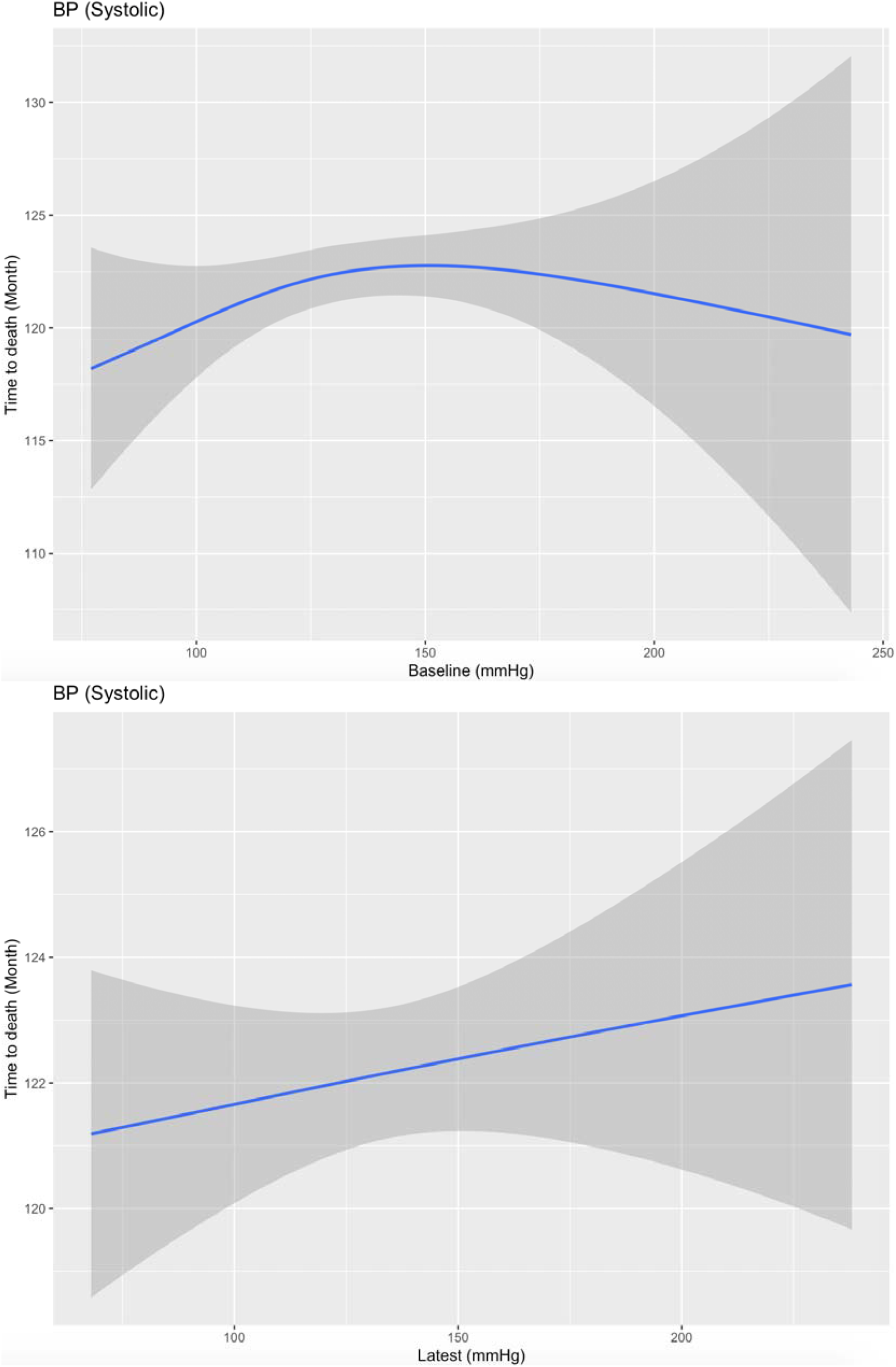
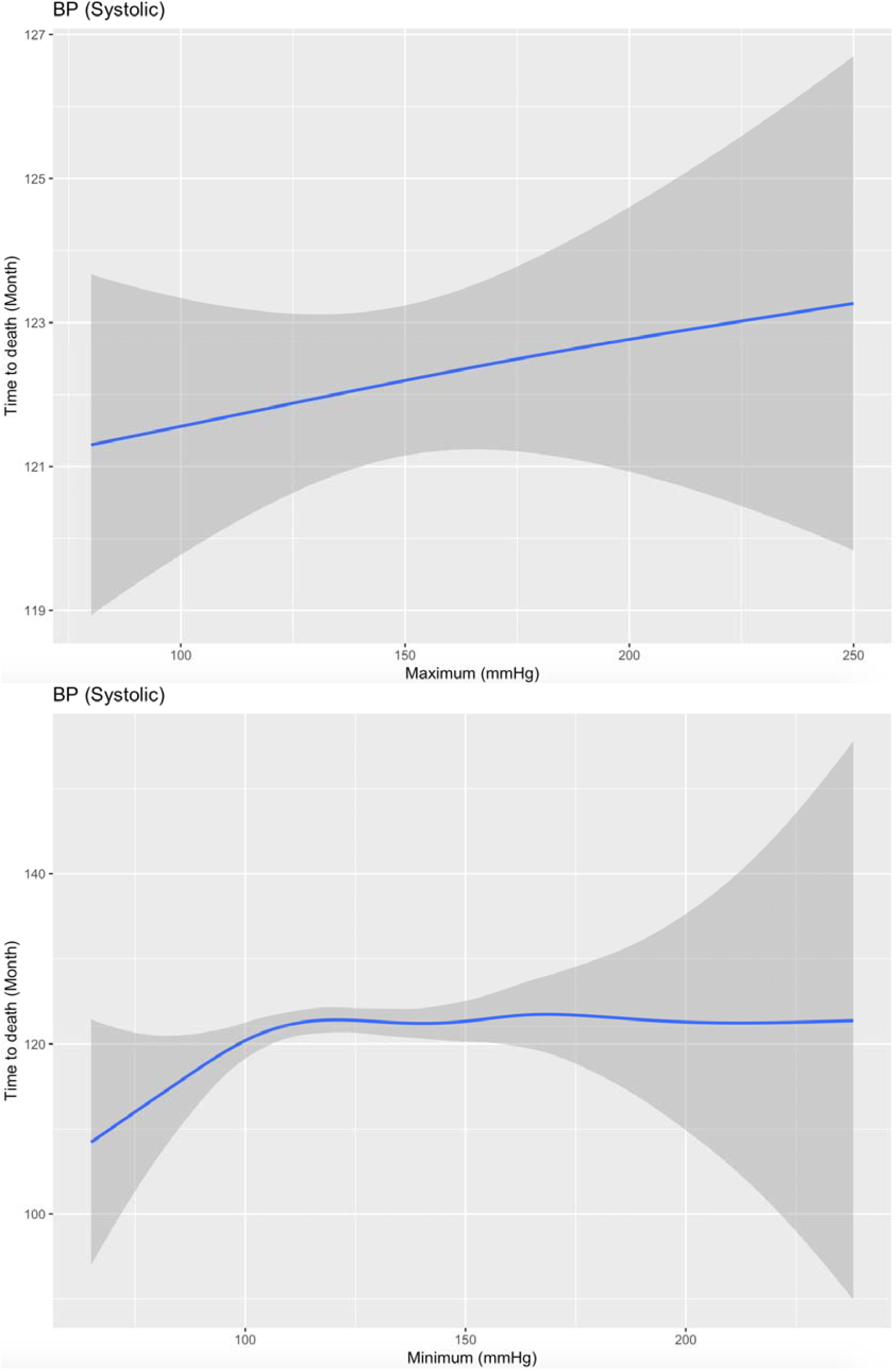
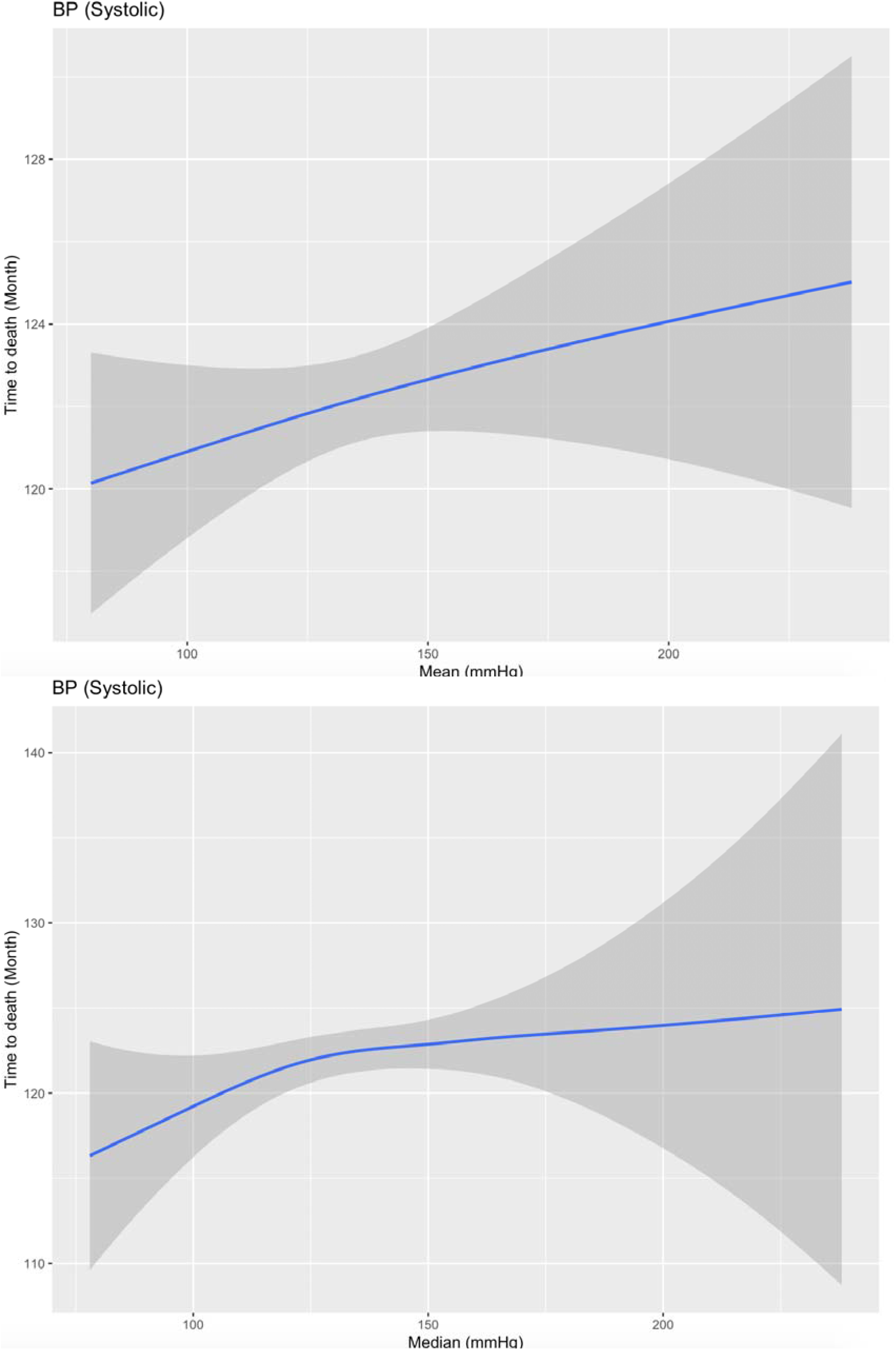
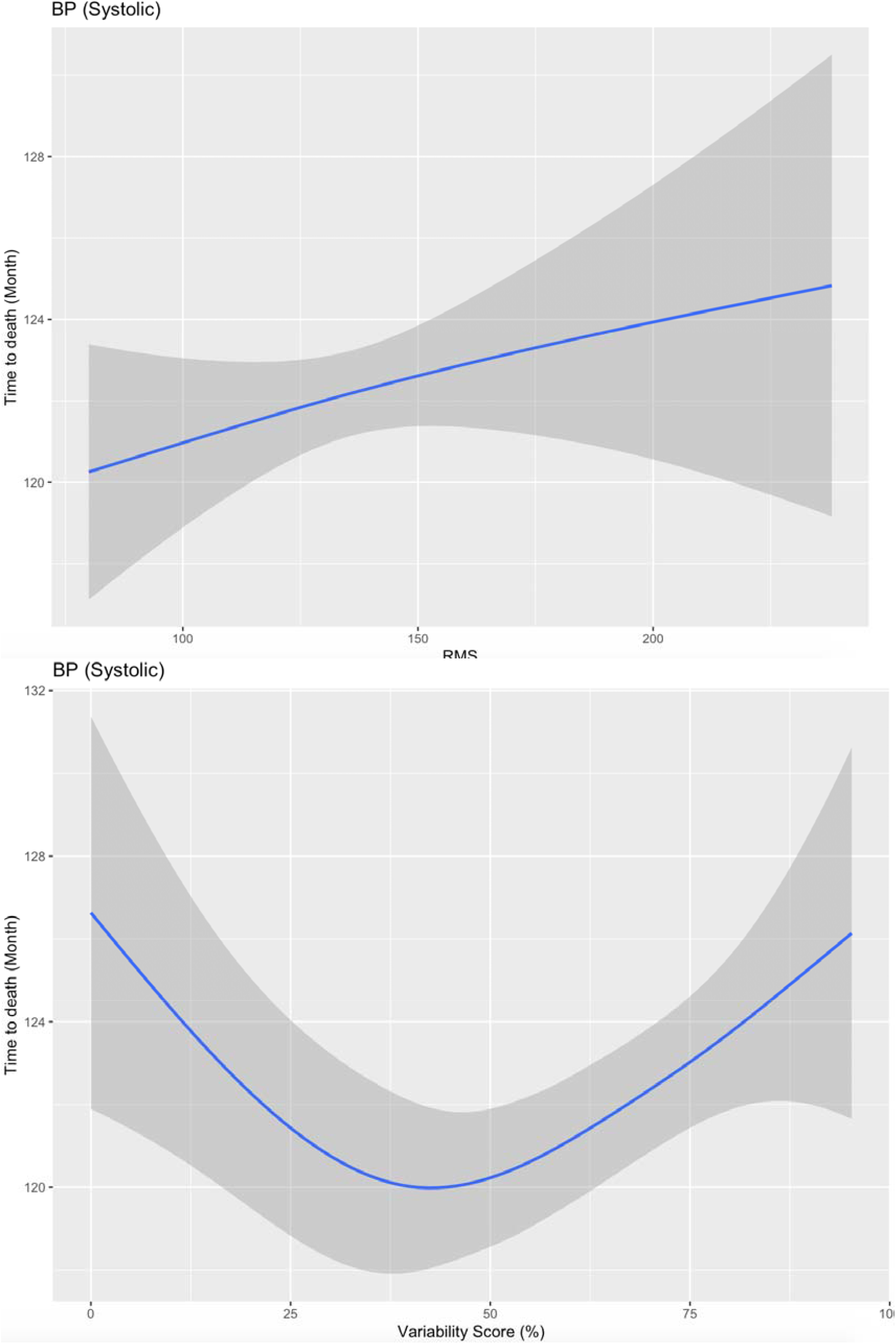
Associations of time-to-death of all-cause mortality with systolic SP and its variability measures.

Laboratory examinations that are significantly predictive for all-cause mortality and adverse cardiovascular outcomes were also identified. In terms of prediction strengths of blood count laboratory examinations for primary and secondary outcomes, higher MCV (HR: 1.019, 95% CI: [1.015,1.023], p<0.001), monocyte (HR: 2.31, 95% CI: [2.09,2.55], p<0.001), neutrophil (HR: 1.10, 95% CI: [1.09,1.11], p<0.001), white blood counts (HR: 1.054, 95% CI: [1.049,1.059], p<0.001), and MCH (HR: 1.03, 95% CI: [1.02,1.04], p<0.001) were significant positive predictors of all-cause mortality. Higher eosinophil (HR: 1.35, 95% CI: [1.15,1.58], p<0.001), monocyte (HR: 1.72, 95% CI: [1.4,2.11], p<0.001), neutrophil (HR: 1.04, 95%CI: [1.02,1.06], p<0.001), and WBC (HR: 1.03, 95% CI: [1.02,1.05], p<0.001) were significant positive predictors of heart failure. Higher MCV (HR: 1.011, 95% CI: [1.006,1.017], p<0.001), monocyte (HR: 1.549, 95% CI: [1.307,1.836], p<0.001), neutrophil level (HR: 1.034, 95% CI: [1.018,1.051], p<0.001), WBC (HR: 1.026, 95% CI: [1.016,1.036], p<0.001) and MCH (HR: 1.026, 95% CI: [1.012,1.041], p<0.001) were significant positive predictors of TIA/stroke.

In terms of liver and renal function tests, higher urate (HR: 18.1, 95% CI: [11.73,27.93], p<0.001), urea (HR: 1.118, 95% CI: [1.114,1.123], p<0.001), creatinine (HR: 1.005, 95% CI: [1.005,1.005], p<0.001), ALP (HR: 1.004, 95% CI: [1.004,1.004], p<0.001), and bilirubin (HR: 1.007, 95% CI: [1.005,1.01], p<0.001) levels were significant positive predictors of all-cause mortality. Higher urate (HR: 25.64, 95% CI: [12.06,54.49], p<0.001), urea (HR: 1.06, 95% CI: [1.05,1.07], p<0.001), and creatinine (HR: 1.003, 95% CI: [1.002,1.003], p<0.001) were significant predictors of heart failure. Higher urea (HR: 1.055, 95% CI: [1.036,1.073], p<0.001) and creatinine (HR: 1.002, 95% CI: [1.001,1.003], p<0.001) levels were significant predictors of acute myocardial infarction. Higher urate (HR: 5.874, 95% CI: [3.117,11.07]), urea (HR: 1.054, 95% CI: [1.045,1.063], p<0.001), and creatinine levels (HR: 1.003, 95% CI: [1.003,1.004], p<0.001) were significant predictors of TIA/stroke.

For diabetes mellitus and lipid tests, higher glucose level (HR: 1.061, 95% CI: [1.055,1.068], p<0.001) was positively predictive for all-cause mortality, and was also predictive for complications of heart failure (HR: 1.047, 95% CI: [1.034,1.061], p<0.001) and TIA/stroke (HR: 1.023, 95% CI: [1.012,1.034], p<0.001).

## Discussion

The major findings of the present study are as follows:

1. Both blood pressure value and its temporal variability can predict mortality and complications in incident heart failure, acute myocardial infarction and TIA/stroke.
2. Generative additive models found the existence of nonlinear inverse U-shaped relationship between baseline/latest/maximum/minimum/mean/median/RMS measures of diastolic blood pressure and time-to-death for all-cause mortality, and low baseline/latest/maximum/minimum/mean/median/RMS systolic blood pressure subgroup are significantly associated with shorter time-to-death for all-cause mortality.
3. Higher variance/SD/CV/variability score of both diastolic systolic blood pressure is associated with increased risks of all-cause mortality, and incident heart failure, acute myocardial infarction and TIA/stroke (HR>1 and p<0.001).
4. Higher level of MCV, monocyte, neutrophil, WBC, MCH, urate, urea, creatinine, ALP level, bilirubin, glucose are significant positive predictors of all-cause mortality. Higher level of eosinophil, monocyte, neutrophil, WBC, urate, urea, creatinine, glucose are significant predictors of heart failure. Higher level of urea and creatinine are significant predictors of acute myocardial infarction. Higher level of MCV, monocyte, neutrophil, WBC, MCH, urate, urea, creatinine, and glucose were significant positive predictors of TIA/stroke.

Previous studies have established a J-shaped relationship between systolic and diastolic blood pressure and all-cause mortality ^7-9^. This study provides further evidence that there is a U-shaped relationship in our territory-wide family medicine cohort. Moreover, visit-to-visit blood pressure variability has been shown to increase non-linearly with all-cause mortality risk in the general population ^10^ and specific disease populations such as diabetes mellitus ^11^, myocardial infarction ^12^ and chronic kidney disease ^13^. Recent work has reported significant associations between higher systolic blood pressure variability in individuals with and without hypertension and increased risks of all-cause mortality, coronary artery disease, stroke, and end-stage renal failure in a large cohort of U.S. veterans ^14^. Furthermore, the present study provides further evidence that different variability measures of systolic and diastolic blood pressure significantly predicted for all-cause mortality and complications of heart failure, acute myocardial infarction, and TIA/stroke in a general family medicine Chinese cohort.

### Limitations

Several limitations of this study should be noted. Firstly, as with other observational studies, this study is limited by potential under-coding of comorbidities, missing data, and coding errors. Secondly, the duration of the complications and the prescribed treatments were not accounted for, which could affect the interpretation of blood pressure value and variability measurements. Thirdly, this study is conducted based on a Hong Kong cohort, and it is expected that external validity through comparisons with studies from other different country of origin reporting the association between blood pressure variability and adverse outcomes could be conducted for further confirmation. Finally, it may well be that the association between blood pressure variability and mortality could be explained by better blood pressure control from treatment. This will need to be examined in future studies by examining the relationship between ongoing treatment and blood pressure values on follow-up beyond the present period over which blood pressure measurements were included.

## Conclusion

Nonlinear inverse U-shaped relationships were observed between blood pressure and its variability measures and all-cause mortality. Higher blood pressure variability was associated with increased risk of all-cause mortality, heart failure, acute myocardial infarction and TIA/stroke.

## Data Availability

Data of this study are deposited in:
https://zenodo.org/record/4383385
They are also uploaded here for academic purposes.

