## Supplementary Appendix for "Associations between Five-Year Blood Pressure Variability and Risk of Cardiovascular Events and Mortality"

**Table 1. Codes for Secondary Complication Outcomes**

| **Comorbidity** | **Diseases and Codes** |
| --- | --- |
| Heart failure | CONGESTIVE HEART FAILURE: 428  HYPERTEN HEART DIS W CHF: 402.91  HEART FAILURE NOS: 428.9  LEFT HEART FAILURE: 428.1  RHEUMATIC HEART DIS NOS: 398.9  CHR PULMON HEART DIS NOS: 416.9  HEART FAILURE: 611.79  OTHER HEART BLOCK: 79.99  CHR PULMON HEART DIS NEC: 617.3  HEART VALVE REPLAC NEC: 621.8  HEART DISEASE NOS: V22.1  RHEUMATIC HEART FAILURE: 478.31  CONG HEART ANOMALY NEC: 891.1  ANEURYSM, HEART (WALL): 225.4  ILL-DEFINED HEART DIS: 211.4  CONG HEART ANOMALY NOS: 57.9  OBSTRUCT HEART ANOM NEC: 389.11  CONGENITAL HEART BLOCK: 303.02  OTH RHEUMATIC HEART DIS: 727.64  FAM HX-ISCHEM HEART DIS: 429.4  ANEURYSM OF HEART NEC: 711.09  CARDIAC DYSRHYTHMIAS NEC: 198.5  CARDIAC ARREST: 427.5  CARDIAC DYSRHYTHMIA NOS: 784.4  CARDIAC DYSRHYTHMIAS: 288.3  CARDIAC SEPTAL DEFECT: 916.9 |
| Myocardial infarction | ACUTE MYOCARDIAL INFARCT: 501  Anterior AMI, initial: 410.11  Latrl AMI NEC, init episd: 410.51  AMI, initial epi: 410.91  INFER AMI NEC-INIT EPISD: 386.11  POST MI SYNDROME: 411  LATRL AMI NEC-INIT EPISD: 830  AMI NOS - EPISODE NOS: 478.7  AMI NEC-INITIAL EPISODE: 758 |
| TIA/stroke | HEAT STROKE & SUNSTROKE: 457.9  CHR ISCHEMIC HRT DIS NEC: 720  OTH CHR ISCHEMIC HRT DIS: 681.1  AC ISCHEMIC HRT DIS NEC: 473.9  OTH AC ISCHEMIC HRT DIS: 290.3  SCRN-ISCHEMIC HEART DIS: 524.02  CHR ISCHEMIC HRT DIS NEC: 720  CHR ISCHEMIC HRT DIS NOS: 414.9  CEREBROVASCULAR ANOMALY: 215.3 |

**Table 2. Codes for Prior Comorbidities**

| **Comorbidity** | **Codes** |
| --- | --- |
| Respiratory | 786.09, 518.81, 780.53, 137, E912, 465.9, 518.81, 518.81, 79.6, 518.81, 519.8, 780.59, 799.1, 780.57, 518.82, 480.1, 786.3, 519.8, 997.3, 165.9, 519.9, 648.91, 162.9, 162.3, 197, 162.5, 162.4, 486, 518.89, 496, 162.8, 415.1, V10.11, 518, 162.9, 11.96, 482.1, 507, 513, 11.9, 511.8, 511.1, 11.94, 516.8, 793.1, 482.4, 507, 515, 197, 11.93, 482, 482.83, 518.4, 482.3, 482.2, 415.1, 502, 518.89, 235.7, 793.1, 934.8, 516.9, 136.3, 38.49, 506, 112.4, 487, 481, 117.9, 38.2, 518, 11.95, 79.89, 518.1, 480.9, 505, 516.8, 495.9, 518.3, 11.23, 416.8, 513, 397.1, 117.3, 483, 508, 998.81, 416, 514, 861.21, 502, 934.8, 480.8, 648.93, 11.2, 492.8, 484.6, 78.5, 484.1, 516.3, 415.1, 416.9, 415, 416.9, 429.89, 415, 747.49, 745, 417, 770.7, 427.5, 416.9, 416, 416.8, 746.02, 573.8, 642.9, 416, 747.3, 747.3, 770.3, 779.8, 515, 424.3, 416, 417.8, 747.3, 747.3, 745.4, 518.81, 786.09, V12.6, 478, 748.5, 162.9, 996.84, 748.5, 748.6, V42.1, 748.5, 11.05, 162, 518, 747.42, 518.89, 748.5, 517.2 |
| Kidney | 198.7, 189, 189, 585.9, V56.0, 189.1, 584.9, 189.1, 593.9, 189, 189, 189.8, 239.5, 189, 583.81, 593.9, 591, V10.52, 250.4, 591, 255.4, 590.8, 586, 585.1, 591, 189, 591, 239.7, 788, 996.39, 189.1, 588.9, 592, 996.39, 250.4, 593.2, 255.4, 572.4, 194, 255, 453.3, 198, 585.9, 996.39, 996.39, 590.1, 591, 591, 223, 591, 584.9, 585.9, 250.41, 996.39, 581.9, 227, 593.5, 583.89, 593.89, 404.93, 255.5, 788.9, 250.43, 227, 405.92, 592, 753.12, 996.39, E879.1, V42.0, 592, 580.89, 403.9, 593.9, V59.4, 585.9, 580.9, 250.41, 996.39, 585.9, 794.4, 584.8, 584.5, 255.9, 441.4, V58.49, 404.11, 593.9, 592, 585, 759.1, 753.11, E879.1, 753.15, 585.9, 274, 588.8, 403.91, 404.9, 404.91, 404.92, 403, 404.01, 779.8, 250.4, 227, 227, 227, 404.93, 996.81, 593.89, 753.8, 593.2, 592, 753, 996.81, 223, 589.1, 996.81, 582.9, 585.9, 996.81, 753.1, 753.3, E878.0, 593.9, 753.3, 753.17, 583.9, 593.9, 589, 866 |
| Endocrine | 202.8, 200.1, 200.12, 201.9, 204, 202.88, 200.18, 196, 204.01, 785.6, 200.11, 200.13, 202.8, 785.6, 202.85, 202.81, 202.8, 202.8, 196, 196.9, 202.8, 202.82, 785.6, 202.8, 202, 202.87, V10.79, 785.6, 202.84, 202.01, 196.8, 457.1, 12.1, 785.6, V10.79, 196.5, 196.2, 238.7, 196.1, 200.14, 457.2, 238.7, V10.61, 201.9, 457, 202.8, 289.3, 245.2, 238.7, 785.6, 457.9, 785.6, 202.93, 196.9, 202.97, 757, V10.71, 288.8, 204.1, 202.83, 457.1, 289.3, 785.6, V77.9, 237.4, 239.7, 198.89, 623.5, 259.9, 200.2, V10.71 |
| Diabetes mellitus | 251.2, 362.01, 362.02, 250.4, 250.82, 790.2, 790.6, 250.5, 250.5, 250.6, 357.2, 790.2, 250, 250.4, 250.6, 250.8, 250.51, 250.5, 250.8, 250.82, 250.51, 250.51, V77.1, 250.12, 251.2, 250.12, 250.5, 250.83, 251.2, 250.41, 251.1, 250.52, 250.5, 648.81, 250.43, 250.53, 250.53, 250.81, 250.22, 250.13, 250.22, 250.83, 250.41, 250.5, 250.52, 250.52, 250.82, 253.5, V18.0, 588.1 |
| Hypertension | 401.9, 401.9, 250.82, 790.6, 401.9, 401.9, 250.82, 796.2, 402.9, 250.83, 405.99, 642.93, 642.01, 642.91, 401.9, E942.6, 405.09, 403.9, 437.2, 401, 401.1, 401, 642.33, 348.2, 779.8, 365.04, 572.3, 416, 416, 405.91, 416.8, 642.3 |
| Gastrointestinal | 153.3, 154.1, 153.9, 569.89, 154, 153.1, 578.9, 560.9, 569.3, 537.89, 558.9, 562.1, 153.6, 239, 532.3, 569.89, 532.7, 535.6, 558.9, 38.42, 569.89, 8.45, 153.2, 569.49, 79.89, 532.9, V58.11, 569, 154.1, 41.4, 537.89, 152.1, 578.9, V10.05, 787.8, 197.4, 535.5, V10.06, 9, 569.83, 569.6, 153.4, 560.9, 537.3, 41.04, 569.84, 239, 569.81, 8.8, 535, 560.9, 532, V45.89, V12.72, 532.4, V10.09, 560.81, 235.2, 38.49, 8.45, 235.2, 532.9, 569.81, 537.89, 557.9, 569.41, 997.4, 14.8, 787.99, 8.46, 535.5, 569.41, 997.4, 578.9, 569.82, 537.9, 560.1, 569.82, 557.9, 211.3, 556.9, 562, 558.9, 578.9, 536.9, 8.46, 535.6, 566, V71.9, 569.49, 564.3, V44.4, 569.89, 564.8, 8.46, 569.83, 997.4, 997.4, 997.4, 562.11, 211.2, 9.1, 211.3, 8.47, 8.5, 211.3, 569.83, 532.1, 535.61, 560, 569.83, 565.1, 619.1, 152.9, 568, 566, 569.43, 152, 8.46, 562.11, 8.61, 569.83, 569.83, 569.81, 596.1, 535.5, 151.4, 151.9, 151.5, 151.8, 151.1, 456.8, 531.7, 535.4, 531.3, V15.2, 537.89, 211.1, 531.9, V10.04, 235.2, 531, 531.4, 456.8, 535.1, 151.3, 230.2, 151.6, 535, 211.1, 536.3, 535, V10.04, 535.51, 578.9, 531.1, 535.1, 456.8, 531.5, 537.84, 535.01, 530.7, 535.1, 535.2, 535.5, 537.6, 202.83, 535.1, 531.4, 558.9, 558.9, 558, 569.85, 153, 555.1, 562.1, 562.13, 562.11, 562.12, 569.83, V76.49, 560.2, 230.4, 569.3, 154 |
